# Increased Risk of Severe COVID-19 Disease in Pregnancy in a Multicenter Propensity Score-Matched Study

**DOI:** 10.1101/2021.06.18.21258899

**Authors:** Liviu Cojocaru, Myint Noe, Autusa PahlavaN, Alissa Werzen, Hyunuk Seung, Young Chae Jessica Yoo, Patricia Tyson, Shivakumar Narayanan, Shifa Turan, Ozhan M. Turan, Joel V. Chua

**Author notes:** <u>Correspondence</u>: Liviu Cojocaru, MD: Division of Maternal-Fetal Medicine. Department of Obstetrics, Gynecology and Reproductive Science, University of Maryland School of Medicine, Baltimore, MD, USA.,.

## Abstract

**Background:** Respiratory infections have long been associated with higher maternal and perinatal morbidity. Early data did not report an increased risk of SARS-CoV-2 infection or disease severity in pregnancy. However, surveillance data from the Center for Disease Control and Prevention (CDC) indicates a higher risk of severe disease and death in pregnant women with symptomatic SARS-CoV-2 infection, although this data is subject to ascertainment bias.

**Objective:** To explore the association between COVID-19 disease severity and pregnancy in our university-based hospital system using measures such as COVID-19 ordinal scale severity score, hospitalization, intensive care unit admission, oxygen supplementation, invasive mechanical ventilation, and death.

**Study design:** We conducted a retrospective, multicenter case-control study to understand the association between COVID-19 disease severity and pregnancy. We reviewed consecutive charts of adult females, ages 18-45, with laboratory-confirmed SARS-CoV-2 infection in six months between March 1, 2020, and August 31, 2020. Cases were patients diagnosed with COVID-19 during pregnancy, whereas controls were not pregnant at the time of COVID-19 diagnosis. Primary endpoints were the COVID-19 severity score at presentation (within four hours) and the nadir of the clinical course. The secondary endpoints were the proportion of patients requiring hospitalization, intensive care unit admission, oxygen supplementation, invasive mechanical ventilation, and death.

**Results:** A higher proportion of pregnant women had moderate to severe COVID-19 disease at the nadir of the clinical course than nonpregnant women (25% vs. 16.1%, *p*=0.04, respectively). While there was a higher rate of hospitalization (25.6% vs. 17.2%), ICU admission (8.9% vs. 4.4%), need for vasoactive substances (5.0% vs. 2.8%), and invasive mechanical ventilation (5.6% vs. 2.8%) in the pregnant group, this difference was not significant after the propensity score matching was applied.

We found a high rate of pregnancy complications in our population (40.7%). The most worrisome is the rate of hypertensive disorders of pregnancy (20.1%).

**Conclusions:** In our propensity score-matched study, COVID-19 in pregnancy is associated with an increased risk of disease severity and an increased risk of pregnancy complications.

## Introduction

Coronavirus disease 2019 (COVID-19), caused by the severe acute respiratory syndrome coronavirus-2 (SARS-CoV-2), exhibits clinical features similar to respiratory viral infections such as influenza, severe acute respiratory syndrome (SARS), and Middle East respiratory syndrome (MERS), that have been associated with higher maternal and perinatal morbidity ^1, 2^. Early data did not support an increased risk of SARS-CoV-2 infection or disease severity in pregnancy^3, 4^. However, more recent data from the Center for Disease Control and Prevention (CDC) indicates a higher risk of severe disease and death in pregnant women with symptomatic COVID-19, although this data was limited by ascertainment bias and other limitations of a surveillance study^5, 6^.

Despite the incongruence between published studies, pregnancy appears to be associated with increased risk of hospitalization, intensive care unit (ICU) admissions, invasive mechanical ventilation (IMV), extracorporeal membranes oxygenation (ECMO), and death ^1, 5, 6^.

Several physiologic and immunologic changes during pregnancy predispose pregnant women to complications related to COVID-19 pneumonia. Increased oxygen consumption (15-20%), low functional residual capacity, and increased renal excretion of bicarbonate to compensate for the respiratory alkalosis result in diminished compensatory homeostatic reserve^7, 8^. Additionally, decreased cell-mediated immunity, helper-T-cell numbers, natural killer activity, along with modulatory effects of T helper 17/regulatory T cells, may result in a decreased immunologic host response ^9-11^. Moreover, during pandemics, the unique needs of pregnant women are sometimes overlooked, whether due to inadequate oxygen supplementation ^12^, decrease in quality of care ^13^, or exclusion from clinical trials^14^. The incidence of COVID-19 is also higher in pregnant women who are Hispanic or non-Hispanic Black. The difference in the prevalence of underlying chronic conditions or social determinants of health may impact the severity of presenting illness or the outcome of infection.

To further explore the association between COVID-19 disease severity and pregnancy, we conducted a retrospective case-control study comparing pregnant and nonpregnant women infected with SARS-CoV-2 in our university-based hospital system. We hypothesized that pregnant women with COVID-19 are at an increased risk of disease severity compared with nonpregnant controls. First, we characterized demographic data, comorbidities, treatments, and outcomes of COVID-19 in pregnant women. Then we aimed to determine whether there is an increased risk of COVID-19 severity in pregnancy using measures such as the need for hospitalization, ICU admission, need for supplemental oxygen, IMV, and death.

## Materials and Methods

We conducted a retrospective, multicenter case-control study to understand the association between COVID-19 disease severity and pregnancy. We reviewed consecutive charts of adult females, ages 18-45, who had laboratory-confirmed SARS-CoV-2 infection in six months between March 1, 2020, and August 31, 2020. Cases were patients diagnosed with COVID-19 during pregnancy, whereas controls were not pregnant at the time of COVID-19 diagnosis. We excluded cases with a compromised immune system (human immunodeficiency virus infection, active malignancy, solid organ or hematopoietic stem cell transplant) and chronic conditions that would require chronic supplemental oxygen therapy. Patients with insufficient data for assessing the COVID-19 diagnosis, pregnancy status, or clinical outcome were also excluded. The Institutional Review Board at the University of Maryland, Baltimore, approved the study under the protocol HP-00093213. The article was prepared following Strengthening the Reporting of Observational Studies in Epidemiology (STROBE) guidelines ^15^. Data were collected and entered into a secure database, Research Electronic Data Capture (REDCap) software^16, 17^, hosted at the University of Maryland, Baltimore.

### COVID-19 Severity Score

The severity of the disease was determined based on and adapted from the National Institute of Allergy and Infectious Disease (NIAID) eight-point ordinal severity score^18^. The lowest score of 0 was given to asymptomatic patients, and the worst score of 7 was assigned to cases of death. A symptomatic patient who did not require hospitalization was given a score of 1 (mild disease), while those who needed hospitalization but did not require oxygen supplementation were given a score of 2 (moderate disease). Patients who required both hospitalization and oxygen supplementation were given either a score of 3 if they required low flow oxygen supplementation (less than six liters) or 4 if they required high flow oxygen supplementation (six or more liters) or non-invasive pressure ventilation. Patients who required invasive mechanical ventilation (IMV) received a score of 5. If, in addition to IMV, patients required blood pressure support with vasoactive substances, renal replacement therapy (RRT), or extracorporeal membranes oxygenation (ECMO), patients received a score of 6. Patients with a COVID-19 severity score of three or higher are considered to have severe disease.

### Study endpoints

Primary endpoints were the COVID-19 ordinal scale score at presentation (within four hours) and the nadir of the clinical course. The secondary endpoints were the proportion of patients requiring hospitalization, oxygen supplementation, ICU admission, IMV, and their respective duration in days. All-cause mortality and COVID-19 related mortality between groups were evaluated as well.

For cases, additional endpoints analyzed were the proportion of patients with adverse maternal outcomes [early pregnancy loss less than 20 weeks (EPL), later pregnancy loss of more than 20 weeks (LPL), neonatal demise (ND), fetal growth restriction (FGR), hypertensive disorders of pregnancy (HDP), preterm delivery (PTD) and gestational diabetes (GDM)].

### Statistical analysis and propensity score matching

Descriptive statistics were used for frequency, median, and mean. We employed propensity score matching (PSM) to lessen the potential influence of confounding factors and increase the reliability of the results. The PSM was performed using age, race/ethnicity, body mass index, and comorbidities (hypertension, diabetes mellitus, heart disease, chronic pulmonary disease) (Table 1). It formed matched sets of cases and controls who share a similar propensity score. Optimal one-to-one matching used the logit of the propensity score as the matching metric. To assess PSM results, the standardized difference for variables included in the propensity score model before and after PSM was provided in Supplementary Figure 1.

**Table 1.** Baseline characteristics of non-propensity matched and propensity-matched populations (1:1).

| Characteristics | Non-Propensity Matched |  |  |  | Propensity Matched* |  |  |  |
| --- | --- | --- | --- | --- | --- | --- | --- | --- |
|  | Total<sup>†</sup><br>(n=1137) | Non-pregnant<br>(n=948) | Pregnant<br>(n=189) | p-value | Total<br>(n=360) | Non-pregnant<br>(n=180) | Pregnant<br>(n=180) | p-value |
| **Age, median (IQR)** | 32.7<br>(25.9, 40) | 33.8<br>(26.8, 40.8) | 28.1<br>(23.8, 34.2) | **<0.001** | 28.7<br>(24, 34.1) | 28.8<br>(23.8, 34.1) | 28.3<br>(24.1, 34) | 0.8 |
| **Race/ ethnicity, n (%)** |  |  |  | **<0.001** |  |  |  | 0.9 |
| Asian, non-Hispanic | 20(1.8) | 17(1.8) | 3(1.6) |  | 6(1.7) | 3(1.7) | 3(1.7) |  |
| Black, non-Hispanic | 568(50.4) | 509(54.2) | 59(31.2) |  | 119(33.1) | 63(35) | 56(31.1) |  |
| White, non-Hispanic | 244(21.6) | 216(23) | 28(14.8) |  | 57(15.8) | 29(16.1) | 28(15.6) |  |
| Hispanic | 249(22.1) | 154(16.4) | 95(50.3) |  | 169(46.9) | 80(44.4) | 89(49.4) |  |
| Other | 47(4.2) | 43(4.6) | 4(2.1) |  | 9(2.5) | 5(2.8) | 4(2.2) |  |
| **BMI, median (IQR)** | 31.1<br>(26, 37.8) | 30.9<br>(25.6, 37.8) | 31.8<br>(27.6, 37.6) | 0.3 | 31.6<br>(26.7, 37.7) | 31.5<br>(25.5, 37.8) | 31.8<br>(27.6, 37.6) | 0.5 |
| **No PMH, n (%)** | 755(66.4) | 613(65) | 142(75.1) | **0.005** | 263(73.1) | 128(71.1) | 135(75) | 0.4 |
| **Lung disease<sup>‡</sup>, n (%)** | 198(17.4) | 173(18.3) | 25(13.2) | 0.1 | 53(14.7) | 30(16.7) | 23(12.8) | 0.3 |
| **Heart disease, n (%)** | 19(1.7) | 17(1.8) | 2(1.1) | 0.8 | 4(1.1) | 2(1.1) | 2(1.1) | 1.0 |
| **HTN, n (%)** | 158(13.9) | 138(14.6) | 20(10.6) | 0.7 | 37(10.3) | 17(9.4) | 20(11.1) | 0.6 |
| **DM, n (%)** | 78(6.9) | 70(7.4) | 8(4.2) | 0.1 | 16(4.4) | 8(4.4) | 8(4.4) | 1.0 |

IQR: interquartile range; n: number; BMI: body mass index; PMH: past medical history; HTN: hypertension; DM: diabetes mellitus.
\*Frequency missing: 9 (race/ethnicity, non-pregnant), 134 (BMI, non-pregnant), 9 (BMI, pregnant) in the non-propensity matched.
<sup>†</sup>Excluded the subjects with missing values (see Figure 1).
<sup>‡</sup>Lung disease: included conditions such as bronchial asthma and chronic obstructive pulmonary disease.

The associations between outcomes or laboratory data and pregnancy were measured using a χ2 or Fisher’s exact test and Wilcoxon rank-sum test. The associations between characteristics and the outcome, COVID-19 score at the nadir of clinical course, were measured using simple logistic regression. For multivariate analysis, a logistic regression model was conducted to evaluate the main exposure and independent factors. The odds ratio (OR) with 95% CI was used to measure the magnitude of the association. The variance inflation factor (VIF) and correlation coefficients were used to identify multicollinearity. The Hosmer-Lemeshow test, the deviance, and Pearson χ2 were used to assure the goodness-of-fit of the logistic model. Analyses were performed with SAS software version 9.4 (SAS Institute, Cary, NC).

## Results

A total of 1,774 patients were identified, of which 637 were excluded due to insufficient information. From the 1,137 patients included in the non-PSM (NPSM) analysis, 189 were pregnant and 948 nonpregnant women. After the PSM algorithm was applied, 180 patients remained in each group. Figure 1 summarizes the study flow.

**Figure 1.**
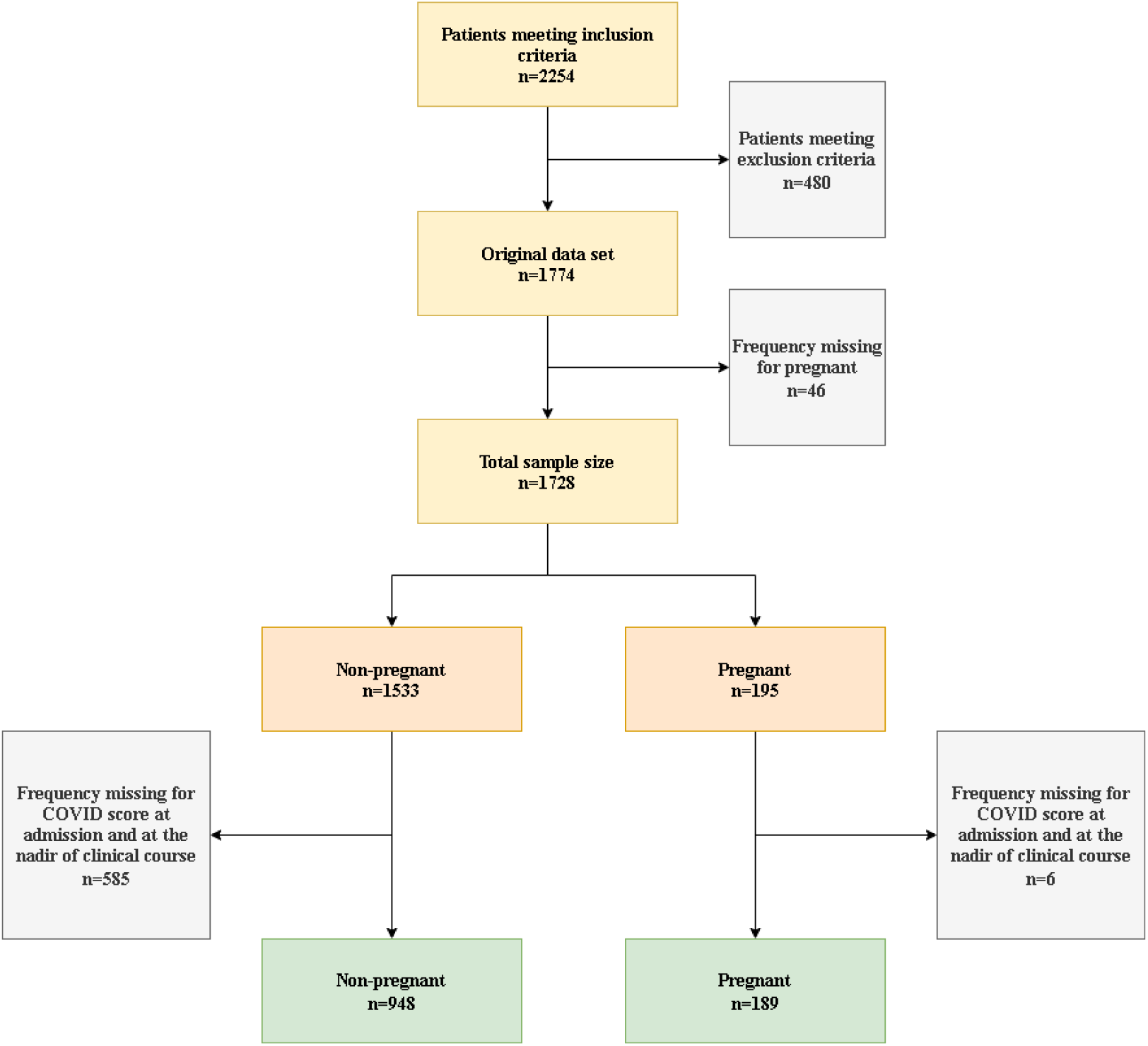
Sampling flow chart and sample size.

### Non-Propensity-Score Match Population

The pregnant group was younger [age 28.1 years (23.8-34.2) vs. 33.8 years (26.8-40.8), *p*<0.001] and had a lower prevalence of preexisting comorbidities (24.9% vs. 35%, *p*=0.005). The ethnic and racial distribution was different between groups, with a higher percentage of Hispanic women (50.3% vs. 16.4%) in the pregnant group and a higher percentage of non-Hispanic Black and non-Hispanic whites (54.2% vs. 31.2% and 23% vs. 14.8%, respectively) in the nonpregnant group, *p*<0.001 (Table 1). The proportion of patients with COVID-19 severity score of 2 or more at presentation and the nadir of the clinical course were higher in the pregnant group (19% vs. 11.8%, *p*=0.007 and 24.9 vs. 15.3%, *p*=0.001, respectively).

Pregnant women had higher rate of hospitalization (25.4% vs. 16%, *p*=0.002), ICU admission (8.5% vs. 3.1%, *p*<0.001), need for vasoactive substances (4.8% vs. 1.8%, *p*=0.03) and IMV (5.3% vs. 1.8%, *p*=0.008) compared to nonpregnant group. There was a trend towards a longer period of need for supplemental oxygen supplementation in the pregnant group vs. nonpregnant group, though this did not reach statistical significance [8 days (4-15) vs. 5 days (3-7), *p*=0.055]. There was no difference in need for renal replacement therapy (0.5% vs 0.5%, *p*=1.0), extracorporeal membrane oxygenation (0.5% vs. 0.2%, *p*=0.4) and death (0.5 vs 0.2%, *p*=0.4) (Table 2).

**Table 2.**
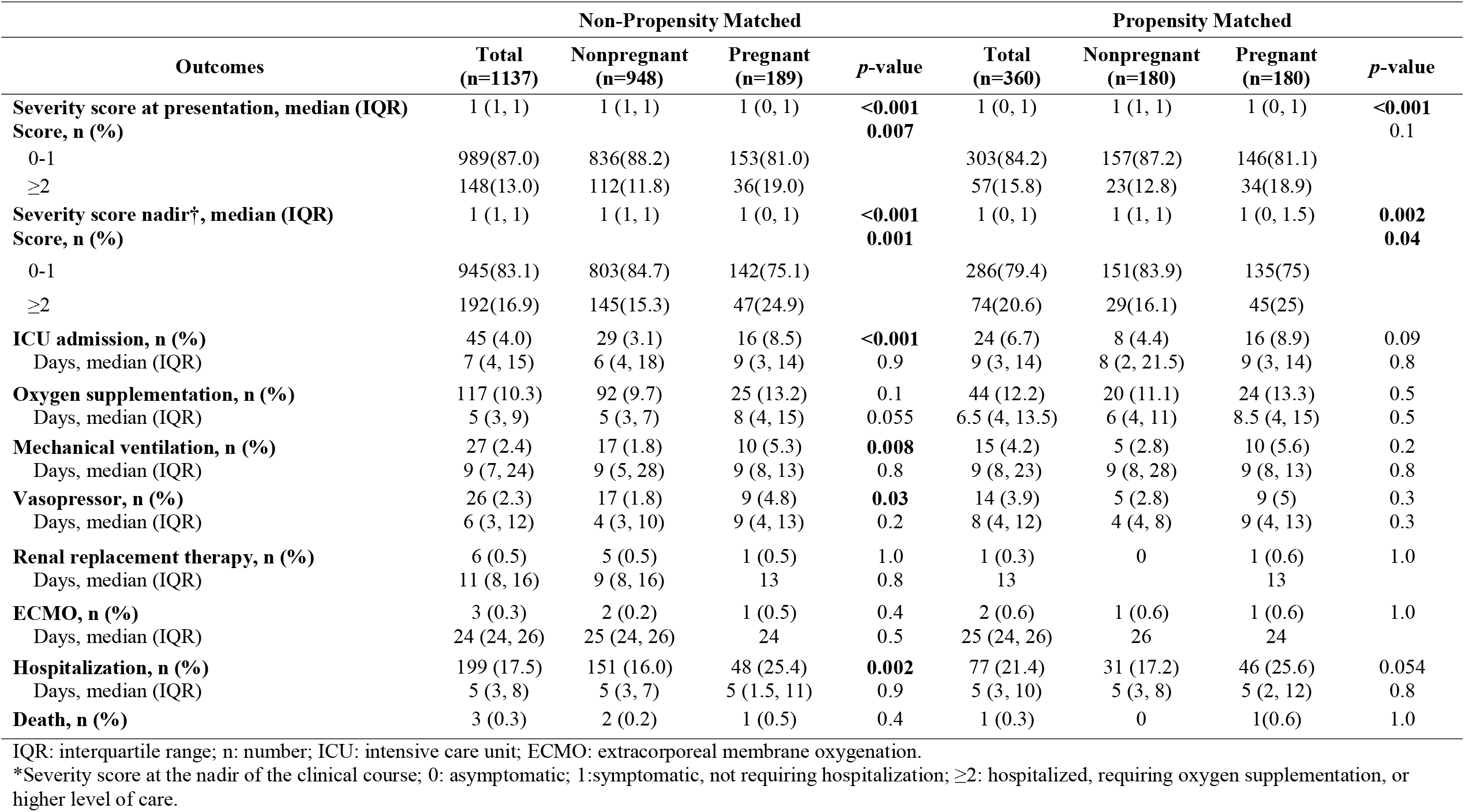
Primary and secondary outcomes in non-propensity matched and propensity-matched populations (1:1).

The review of laboratory analysis revealed that pregnant women had higher leukocyte count (7.4 K/mcl vs. 5.5 K/mcl, *p*<0.001), neutrophils count (5.25 K/mcl vs. 3.45 K/mcl, *p*<0.001), D-dimers values (1,500 ng/mL vs. 600 ng/mL, *p*<0.001) and lower hemoglobin (11.6 g/dL vs. 12.6 g/dL, *p*<0.001), ferritin (54.3 ng/mL vs. 123 ng/mL, *p*<0.001), and serum creatinine (0.5 mg/dL vs. 0.7 mg/dL, *p*<0.001) compared to nonpregnant women (Supplementary Table 3). Also, pregnant women were more likely to have chest x-ray (CXR) findings consistent with COVID-19 (63.3% vs. 36.6%, *p*=0.001) at presentation. There was a trend towards a lower platelet count (220,000/mcl vs. 232,000/mcl, *p*=0.058) and no difference in the lymphocyte count, liver enzyme elevation, total bilirubin, or C-reactive protein. The frequency of abnormal computerized tomography (CT) scan findings was not different between the groups (Supplementary Table 3).

#### Multivariate analysis

The variance inflation factor (VIF) was calculated for multiple regression to determine the independent risk factors (Supplementary Table 2). There were no VIF greater than 2 except no preexisting comorbidities (VIF=4.5), which was excluded from the final model. The backward selection was applied from variables in Supplementary Table 2 (except for no preexisting medical conditions). The overall model was statistically significant likelihood χ2 (7) = 109 with a *p*<0.001 and pseudo *R*^2^ of 17.6%. The Hosmer and Lemeshow χ2 (8) statistic were 8.9, with a p-value of 0.4. Deviance and Pearson goodness-of-fit tests with *p*-values of 1.0 and 0.94. All these goodness-of-fit tests denote that the fitted model is correct. We found that pregnancy, age, Hispanic race, BMI, lung disease, and diabetes are independent risk factors for COVID-19 severity (Table 3). Pregnant women were more likely to have severe disease [OR 2.02 (1.26-3.23), *p*=0.003]. When compared to the white race, the Hispanic population was more likely to have a severe disease [OR=2.86 (1.65-4.94), *p*<0.001]. With every year increase in age and with every unit increase in BMI, there was an increased risk for severe disease [OR 1.08 (1.04-1.11), *p*<0.001, and OR=1.03, (1.009-1.05), *p*=0.004 respectively]. Patients with chronic lung disease and diabetes were more likely to have severe COVID-19 disease [OR 2.31, (1.50-3.57), *p*<0.001, and OR 2.36, (1.34-4.15), *p*=0.003, respectively].

**Table 3.** Multivariate analysis of the COVID-19 severity score* in non-propensity matched and propensity-matched populations (1:1).

| Characteristics | Severity Score*<br>Reference = 0 to 1 | Non-Propensity Matched |  | Propensity Matched |  |
| --- | --- | --- | --- | --- | --- |
|  |  | Adjusted OR<br>(95% CI) | *p*-value | Adjusted OR<br>(95% CI) | *p*-value |
| **Pregnancy** |  |  |  |  |  |
| Yes vs. no | ≥2 | 2.02(1.26, 3.23) | **0.003** | 2.05(1.15, 3.66) | **0.02** |
| **Age** | ≥2 | 1.08(1.04,1.11) | **<0.001** | 1.06(1.01, 1.11) | **0.01** |
| **Race/ ethnicity** |  |  |  |  |  |
| Hispanic vs. White | ≥2 | 2.86(1.65, 4.94) | **<0.001** | 3.38(1.24, 9.15) | **0.02** |
| **Body mass index** | ≥2 | 1.03(1.009, 1.05) | **0.004** | 1.04(1.009, 1.08) | **0.01** |
| **Lung disease<sup>†</sup>** |  |  |  |  |  |
| Yes vs. no | ≥2 | 2.31(1.50, 3.57) | **<0.001** | 4.0(1.89, 8.47) | **<0.001** |
| **Diabetes mellitus** |  |  |  |  |  |
| Yes vs. no | ≥2 | 2.36(1.34, 4.15) | **0.003** |  |  |

IQR: interquartile range; n: number; OR: odds ratio; CI: confidence interval.
\*Severity score at the nadir of the clinical course; 0: asymptomatic; 1:symptomatic, not requiring hospitalization; ≥2: hospitalized, requiring oxygen supplementation, or higher level of care.
<sup>†</sup>Lung disease: included conditions such as bronchial asthma and chronic obstructive pulmonary disease.

**Table 4.** Maternal outcomes in the pregnant population.

| **Maternal outcomes** |  |
| --- | --- |
| **Pregnant complications (overall), n (%)** | 77 (40.7) |
| **Early pregnancy loss, n (%)** | 4 (2.1) |
| **Late pregnancy loss, n (%)** | 3 (1.6) |
| **Neonatal demise, n (%)** | 0 |
| **Fetal growth restriction, n (%)** | 6 (3.2) |
| **Hypertensive disorders of pregnancy, n (%)** | 38 (20.1) |
| **Preterm delivery, n (%)** | 20 (10.6) |
| **Gestational diabetes mellitus, n (%)** | 21 (11.1) |
| **Gestational age\*, median (IQR)** | 222.5 (165, 268) |

IQR: interquartile range; n: number.
\*In days.

### Propensity-Score Matched Population

The matched sets of the pregnant and nonpregnant population had similar age, race/ethnicity, body mass index, and preexisting medical comorbidities. The proportion of patients with COVID-19 severity score of 2 or more was similar at presentation but higher at the nadir of clinical course in the pregnancy group (18.9% vs.12.8%, *p*=0.1, and 25 vs. 16.1%, *p*=0.04, respectively). There was a trend for a higher hospitalization rate in the pregnant group (25.6% vs. 17.2%, *p*=0.054). While the odds ratio for ICU admission and IMV was double in the pregnancy group, this was not statistically significant (8.9% vs. 4.4%, *p*=0.09 and 5.6% vs. 2.8%, *p*=0.2, respectively). There was no observed difference between groups with regards to need for oxygen supplementation (13.3% vs. 11.1%, *p*=0.5), renal replacement therapy (0.6% vs. 0%, p=1), extracorporeal membrane oxygenation (0.6% vs. 0.6%, *p*=1) and death (0.6% vs. 0%, *p*=1) (Table 2).

The PSM did not change the laboratory analysis results, except for lower alanine transferase levels in the pregnant group (20 units/L vs. 30 units/L, *p*=0.004).

Supplementary Table 2 summarizes selected COVID-19 related treatments. Except for the use of corticosteroids, there was no difference in the medical treatment between the pregnant and nonpregnant groups after the PSM was applied.

#### Multivariate analysis

The variance inflation factor (VIF) was calculated for multiple regression to determine the independent risk factors (Supplementary Table 3). There were no VIF greater than 2 except no preexisting comorbidities (VIF=6.1), which was excluded from the final model. The backward selection was applied. The overall model was statistically significant likelihood χ2 (6) = 43 with a *p*<0.0001 and pseudo *R*^2^ of 18.2%. The Hosmer and Lemeshow χ2 (8) statistic were 8, with a *p*-value of 0.4. Deviance and Pearson goodness-of-fit tests with *p*-values of 0.9 and 0.8. All these goodness-of-fit tests denote that the fitted model is correct.

Comparably to the non-propensity-matched population, we found that pregnancy, age, Hispanic ethnicity, BMI, and chronic lung disease are independent risk factors (Table 3). Pregnant women were more likely to have severe disease [OR 2.05 (1.15-3.66), *p*=0.02], Hispanic population being disproportionally affected [OR 3.38 (1.24-9.15), *p*=0.02]. Likewise, with every year increase in age and with every unit increase in BMI, there was an increased risk for severe disease [OR 1.06 (1.01-1.11), *p*=0.001, and OR 1.04 (1.009-1.08), *p*=0.01, respectively]. Additionally, the patients with chronic lung disease and diabetes were more likely to have severe disease [OR 4 (1.89-8.47), *p*<0.001].

### Pregnancy Complications

Slightly above 40% of pregnant women in our study experienced pregnancy complications. The median gestational age at delivery was 32 weeks. Hypertensive disorders of pregnancy were the most common complication (20.1%), followed by preterm delivery (10.6%), gestational diabetes mellitus (11.1%), and fetal growth restriction (3.2%). The early pregnancy loss rate was 2.2%, and the late pregnancy loss 1.6%. There were no cases of neonatal demise.

## Discussion

### Principal findings

Pregnancy was associated with an increased risk of COVID-19 severity at the nadir of clinical course compared to nonpregnant patients. This was statistically significant even after adjusting for baseline demographics and preexisting medical conditions. While there was a higher rate of hospitalization, ICU admission, need for vasoactive substances, and IMV in the pregnant group, this difference was not statistically significant after the propensity score matching was applied. It is possible that this was due to the reduction in the sample size once the propensity score matching was applied, as there was still a trend for higher rates of hospitalization, ICU admission, and IMV in the pregnant group. Nonetheless, pregnancy showed an increased risk of COVID-19 severity compared with a control group with well-balanced demographic and preexisting medical condition characteristics.

In congruence with other studies, the Hispanic population appears to be disproportionately affected. We found a high rate of pregnancy complications in our population (40.7%). The most concerning is the rate of hypertensive disorders of pregnancy (20.1%). While we do not have a pregnant COVID-19 negative control group, the rate of HDP is significantly higher when compared to historical rates (6.9%) from the same state^19^.

### Results in the Context of What is Known

Our data is in congruence with the CDC report and supports that pregnant women are at an increased risk of severe COVID-19. As previously mentioned, several physiologic^7, 8^, and immunologic changes^9-11^ in pregnancy could contribute to an increased risk of respiratory infection severity. However, it is not clear if the increased risk of HDP is a consequence of COVID-19 infection or if these patients are predisposed to both conditions. A higher cytokine levels that are seen with both HDP^20^ and COVID-19^21^, as well as different susceptibility to both conditions with angiotensin converting enzyme 2 polymorphism, might be in favor of the latter.

### Clinical Implications

Pregnant women are at an increased risk of COVID-19 disease severity. Similarly, age, increased BMI, Hispanic ethnicity, and chronic lung disease are independent risk factors. Though, based on the confidence interval close to 1, the size effect of age and BMI may be small. We recommend close surveillance of symptomatic pregnant women with COVID-19, especially in the presence of other risk factors such as Hispanic ethnicity and chronic lung disease. Moreover, we need to prioritize resources and ensure access to new treatment strategies and preventive measures (e.g., vaccination) in this population.

### Research Implications

One lesson learned during the current and previous pandemics is that despite an increased risk of disease severity of infectious diseases in pregnancy, there is a lack of an equitable response in drug development, vaccine studies, enrollment in clinical trial or development of monitoring and management protocols that meet the unique needs of pregnant women^14, 22^. These deficiencies, along with the increased risk of disease severity in pregnancy, further increase the risk of pregnancy outcomes. We should advocate for pregnant women to determine their eligibility and entry into research studies and contribute to the development of pregnancy-specific guidelines. Treatment and vaccine studies should include pregnant women early in the trials as opposed to in the subsequent phases. In addition, further research is required to elucidate the relationship between COVID-19 in pregnancy and pregnancy complications.

### Strengths and Limitations

To our knowledge, this is the first study to evaluated pregnancy impact on COVID-19 severity using a PSM control. Our study’s strengths are a relatively large sample size for both pregnant and nonpregnant groups and a well-matched propensity score population, which decreases the impact of confounding factors. Lastly, our population is predominantly Hispanic and non-Hispanic Black, which has been disproportionately affected by COVID-19^6^ and contributes to understanding risk factors and outcomes in this population.

The limitations of our study are mainly related to the retrospective design. A certain proportion of patients were excluded by design from the data analysis for PSM. Since these patients would have belonged to the nonpregnant control group, this exclusion is unlikely to have influenced the PSM results. There might have been differential testing practices or test availability between pregnant and nonpregnant populations or disadvantaged groups. Even though we excluded non-COVID-19 related hospitalizations, the threshold for hospitalization might have been lower in the pregnant group. Likewise, the pregnancy goal for oxygen saturation of greater than 94% might have resulted in lower thresholds for oxygen supplementation.

Lastly, the study is likely underpowered to detect a difference in rare events such as the need for renal replacement therapy, extracorporeal membrane oxygenation, and death. In addition, while we controlled for the most common comorbidities, the sample size of the PSM group was prohibitive in terms of performing the PSM for all comorbidities. Therefore, our study results may not be generalizable to other populations.

## Conclusion

In our propensity score-matched study, COVID-19 in pregnancy is associated with an increased risk of disease severity and an increased risk of perinatal complications.

## Data Availability

Data available upon a reasonable request.

## Acknowledgments

none.

## Tables, Figures, and Supplementary Tables

**Supplementary Table 1.** Symptoms and laboratory data in non-propensity matched and propensity-matched populations (1:1).

| Outcomes | Non-Propensity Matched |  |  |  | Propensity Matched |  |  |  |
| --- | --- | --- | --- | --- | --- | --- | --- | --- |
|  | Total<br>(n=1137) | Nonpregnant<br>(n=948) | Pregnant<br>(n=189) | *p*-value | Total<br>(n=360) | Nonpregnant<br>(n=180) | Pregnant<br>(n=180) | *p*-value |
| Symptoms, n(%) | 989 (87.0) | 875 (92.3) | 114(60.3) | **<0.001** | 271(75.3) | 163(90.6) | 108(60) | **<0.001** |
| White blood cell<br>count, median (IQR) | n=498<br>6.0 (4.6, 7.9) | n=359<br>5.5 (4.2, 7.2) | n=139<br>7.4 (5.9, 10.2) | **<0.001** | n=201<br>7 (5.3, 9.2) | n=71<br>5.5 (4.1, 8) | n=130<br>7.4 (5.9, 10.1) | **<0.001** |
| Hemoglobin, median<br>(IQR) | n=498<br>12.4 (11, 13.3) | n=359<br>12.6 (11.4, 13.5) | n=139<br>11.6<br>(10.7, 12.5) | **<0.001** | n=201<br>11.9 (10.7, 12.9) | n=71<br>12.5 (10.3, 13.6) | n=130<br>11.7 (10.7, 12.5) | **0.01** |
| Platelet count,<br>median (IQR) | n=498<br>227 (185, 283) | n=359<br>232 (186, 287) | n=139<br>220<br>(176, 270) | 0.058 | n=201<br>224 (184, 275) | n=71<br>236 (196, 298) | n=130<br>219 (176, 267) | 0.06 |
| Neutrophils, median<br>(IQR) | n=443<br>3.8(2.7, 5.8) | n=342<br>3.4 (2.4, 5.2) | n=101<br>5.2 (4.1, 8.0) | **<0.001** | n=163<br>4.7 (3.1, 6.6) | n=68<br>3.5 (2.3, 5.3) | n=95<br>5.2 (4.1, 7.6) | **<0.001** |
| Lymphocytes,<br>median (IQR) | n=438<br>1.4(0.9, 1.83) | n=337<br>1.4 (0.9, 1.9) | n=101<br>1.3 (0.9, 1.8) | 0.5 | n=163<br>1.4 (0.9, 1.9) | n=68<br>1.4 (0.9, 2.0) | n=95<br>1.3 (0.9, 1.8) | 0.4 |
| Creatinine, median<br>(IQR) | n=465<br>0.65 (0.5, 0.8) | n=360<br>0.7 (0.6, 0.83) | n=105<br>0.5 (0.4, 0.6) | **<0.001** | n=170<br>0.56 (0.47, 0.69) | n=72<br>0.63 (0.55, 0.77) | n=98<br>0.5 (0.4, 0.6) | **<0.001** |
| AST, median(IQR) | n=442<br>30 (23, 42) | n=342<br>31 (23, 42) | n=100<br>30 (23, 42) | 0.9 | n=163<br>31 (24, 42) | n=69<br>34 (24, 43) | n=94<br>30 (23, 42) | 0.1 |
| ALT, median(IQR) | n=445<br>21 (14, 37) | n=344<br>22 (15, 39) | n=101<br>20 (14, 35) | 0.4 | n=164<br>24 (14, 46) | n=69<br>30 (18, 51) | n=95<br>20 (14, 33) | **0.004** |
| Total bilirubin,<br>median(IQR) | n=440<br>0.4 (0.3, 0.6) | n=340<br>0.4 (0.3, 0.6) | n=100<br>0.4 (0.3, 0.66) | 0.1 | n=160<br>0.4 (0.3, 0.6) | n=66<br>0.4 (0.3, 0.5) | n=94<br>0.4 (0.3, 0.6) | 0.2 |
| CRP, median(IQR) | n=166<br>4.7 (1.8, 7.8) | n=129<br>4.7 (1.9, 7.8) | n=37<br>4.1 (1.6, 7.7) | 0.9 | n=61<br>4.8 (1.7, 7.6) | n=25<br>5.6 (1.8, 7.1) | n=36<br>4.3 (1.6, 7.8) | 0.9 |
| D-dimers,<br>median(IQR) | n=185<br>709 (430, 1250) | n=153<br>600 (387, 1020) | n=32<br>1500 (984, 2030) | **<0.001** | n=60<br>1009 (610, 1805) | n=30<br>648 (363, 1000) | n=30<br>1505 (1018, 2068) | **<0.001** |
| Ferritin,<br>median(IQR) | n=160<br>99.9 (45.8, 217.1) | n=121<br>123 (56, 262) | n=39<br>54.3(22.6, 73.9) | **<0.001** | n=65<br>61.6 (34.3, 117) | n=27<br>113 (53, 296) | n=38<br>54.7(22.6, 73.9) | **0.003** |
| Abnormal CXR,<br>n(%) | n=443<br>175(39.5) | n=394<br>144(36.6) | n=49<br>31(63.3) | **0.001** | n=121<br>61(50.4) | n=74<br>32(43.2) | n=47<br>29(61.7) | **0.048** |
| Abnormal CT scan,<br>n(%) | n=109<br>84(77.1) | n=98<br>74(75.5) | n=11<br>10(90.9) | 0.5 | n=30<br>26(86.7) | n=20<br>17(85) | n=10<br>9(90) | 0.7 |

IQR: interquartile range; n: number.
ALT: alanine aminotransferase; AST: aspartate aminotransferase; CRP: C-reactive protein; CXR: chest x-ray; CT: computer tomography.

**Supplementary Table 2.**
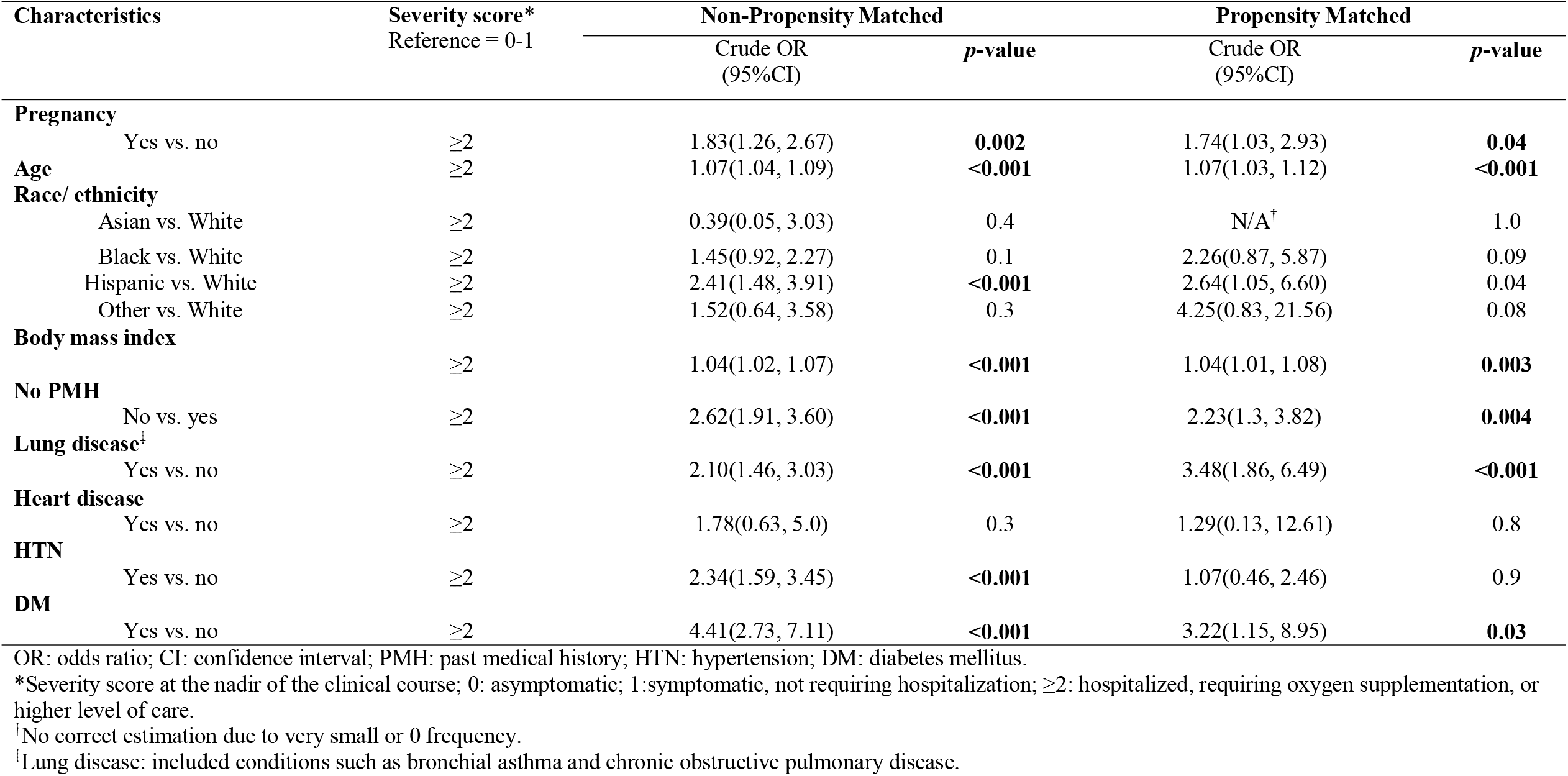
Bivariate analysis of COVID-19 severity score* in non-propensity matched and propensity-matched characteristics (1:1).

**Supplementary Table 3.** COVID-19 related treatments in non-propensity matched and propensity-matched populations (1:1).

| Treatments | Non-Propensity Matched |  |  |  | Propensity Matched |  |  |  |
| --- | --- | --- | --- | --- | --- | --- | --- | --- |
|  | Total<br>(n=1137) | Nonpregnant<br>(n=948) | Pregnant<br>(n=189) | p-value | Total<br>(n=360) | Nonpregnant<br>(n=180) | Pregnant<br>(n=180) | p-value |
| Treatment, n (%) | 131 (11.5) | 102 (10.8) | 29 (15.3) | 0.07 | 50 (13.9) | 23 (12.8) | 27 (15) | 0.5 |
| Remdesivir, n (%) | 43 (3.8) | 34 (3.6) | 9 (4.8) | 0.4 | 19 (5.3) | 11 (6.1) | 8 (4.4) | 0.5 |
| Tocilizumab, n (%) | 9 (0.8) | 9 (1.0) | 0 | 0.4 | 3 (0.8) | 3 (1.7) | 0 | 0.2 |
| Convalescent plasma, n (%) | 9 (0.8) | 7 (0.7) | 2 (1.1) | 0.6 | 4 (1.1) | 2 (1.1) | 2 (1.1) | 1.0 |
| Corticosteroids*, n (%) | 77 (6.8) | 49 (5.2) | 28 (14.8) | **<0.001** | 38 (10.6) | 12 (6.7) | 26 (14.4) | **0.02** |
| Hydroxychloroquine, n (%) | 45 (4.0) | 37 (3.9) | 8 (4.2) | 0.8 | 15 (4.2) | 8 (4.4) | 7 (3.9) | 0.8 |

\*Includes corticosteroids received primarily for fetal lung maturation.

**Supplementary Figure 1.**
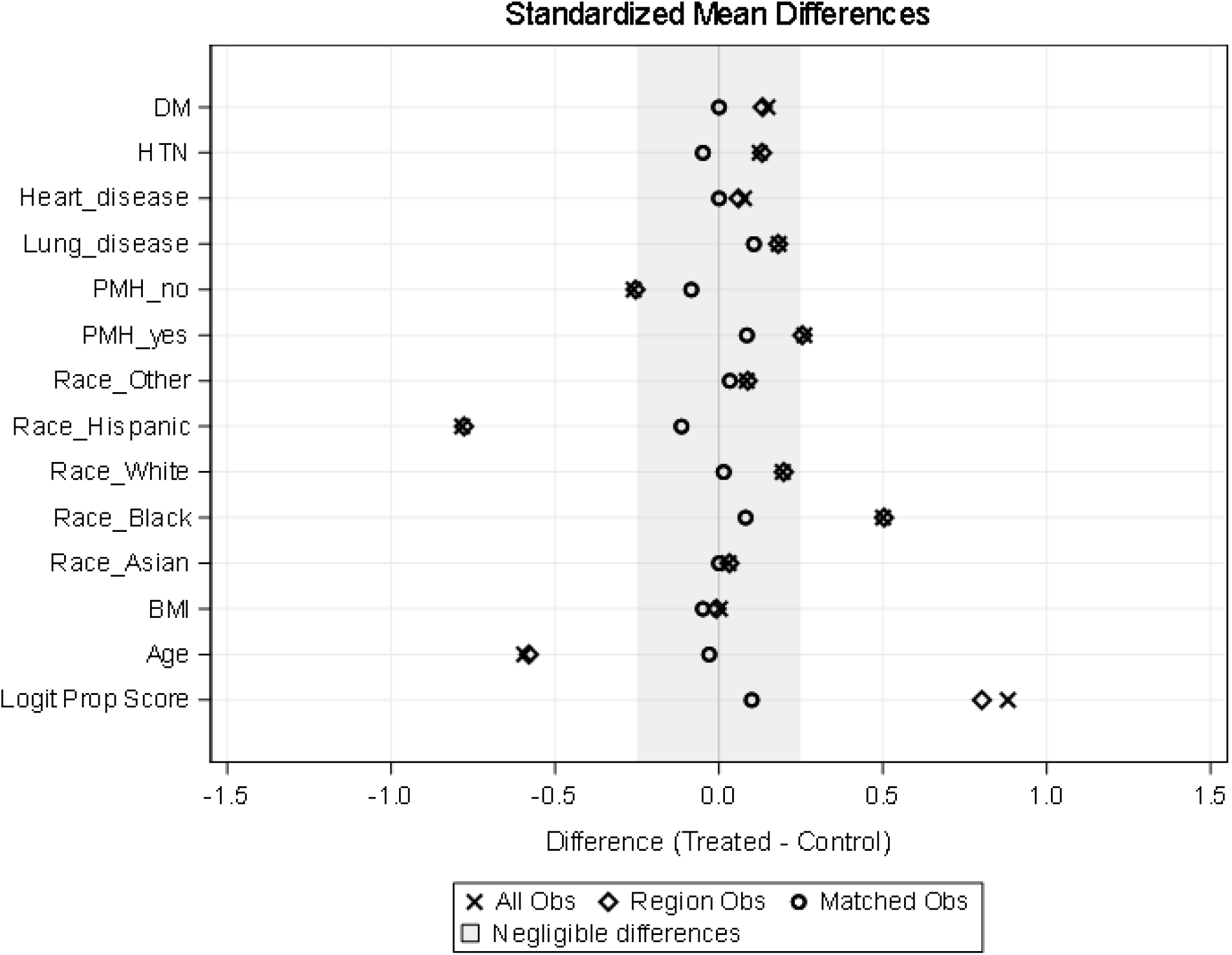
Standardized difference for variables included in the propensity score model before and after propensity score matching. Treated = Pregnant; Control = Nonpregnant. BMI: body mass index; DM: diabetes mellitus; HTN: hypertension; PMH: past medical history.

